# Institutional Standing and Trainee Outcomes in the 2025 US Residency Match

**DOI:** 10.64898/2026.07.09.26357696

**Authors:** Joseph I. Turner, Ariel Arias, Jesse Burk-Rafel, Eric K. Oermann

**Affiliations:** NYU Grossman School of Medicine, New York, New York, USA; Department of Neurological Surgery, NYU Langone Health, New York, New York, USA; Institute for Innovations in Medical Education, NYU Grossman School of Medicine, New York, New York, USA; Global AI Frontier Lab, New York University, New York, New York, USA; Department of Radiology, NYU Langone Health, New York, New York, USA; Center for Data Science, New York University, New York, New York, USA; Neuroscience Institute, NYU Langone Health, New York, New York, USA

**Author notes:** Correspondence: Joseph I. Turner, NYU Grossman School of Medicine, 550 First Avenue, New York, NY 10016, USA.

## Abstract

**Importance:** The transition from medical school to residency forms a national training network, yet its large-scale structure and implications for trainee outcomes remain poorly characterized.

**Objective:** To evaluate the US residency match as a network and assess how institutional position relates to residency placement, educational debt, and specialty choice.

**Design:** Cross-sectional analysis of publicly reported 2025 residency match outcomes.

**Setting:** 107 US MD-granting medical schools and 301 residency institutions with available match data.

**Participants:** 14,616 US MD students matching into residency in 2025 (convenience sample).

**Exposure:** Institutional position within the residency match network, quantified using PageRank network centrality. The relative strength of each school’s graduating class was defined as the median centrality of residency destinations across graduates (placement score).

**Main Outcomes and Measures:** Residency placement outcomes, mean medical school debt at graduation, and specialty choice (primary care vs surgical specialties) in relation to institutional position within the residency match network. Network-derived measures were also compared with NIH funding, residency reputation, and student selectivity.

**Results:** Among 14,616 US MD students matched across 107 medical schools and 301 residency institutions (approximately 73.5% of total US MD cohort), network-derived measures of institutional influence closely aligned with benchmarks of institutional standing such as NIH funding, residency reputation, and student selectivity (Spearman ρ = 0.72–0.86; all p < .001). Graduate outcomes varied systematically across institutions. Graduates of highly connected medical schools were more likely to match into highly connected residency programs (87.3% for top-quintile vs 41.0% for bottom-quintile schools). Schools with higher placement scores had graduates with lower educational debt, reduced entry into primary care, and increased entry into surgical or competitive specialties. Compared with bottom-decile schools, top-decile schools (stratified by placement score) had 37% lower mean graduate debt, 24% lower primary care entry, and 75% higher surgical specialty entry. Higher educational debt was not associated with entry into higher-compensated specialties.

**Conclusions and Relevance:** The residency match network reflects a hierarchical structure of institutional standing. Graduates of higher- and lower-positioned medical schools experience systematically different residency placement outcomes. These findings provide a population-level, behavior-based perspective on institutional influence and its relationship to training pathways.

**KEY POINTS:** *Question:* How does institutional standing relate to residency placement, debt burden, and specialty choice?

*Findings:* In this cross-sectional analysis of 14,616 US MD students entering residency, institutional influence derived from match data aligned with traditional metrics like NIH funding. Graduates of highly connected medical schools matched into highly connected residency programs. These well-connected schools also showed lower student debt, less primary care entry, and higher placement into surgical fields. Higher average debt was not associated with greater entry into higher-paying specialties.

*Meaning:* The residency match operates as a hierarchy, linking a medical school’s position to career pathways and graduate outcomes.

## INTRODUCTION

The transition from medical school to residency in the United States is governed by a centralized matching system that differs from most professional labor markets. Graduating students and residency programs submit ranked preference lists, and placements are determined through a stable matching algorithm used by the National Resident Matching Program (NRMP). This Roth-Peranson algorithm underlies the modern Match^1,2^, and comparable systems are used in specialties such as ophthalmology^3^ and urology^4^. Because the Match produces a single binding placement for each applicant, it generates a large national set of school-to-residency transitions. In 2025 alone, 19,044 U.S. MD seniors matched into PGY-1 positions through 5,773 residency programs^5^. Yet despite the scale of this system, most publicly available analyses remain aggregate and do not directly examine flows between named medical schools and residency destinations.

A network framework is well suited to this problem. Each match outcome can be represented as a directed edge from a student’s home medical school to a residency institution, allowing the Match to be studied as a national training network. In directed networks, PageRank provides a recursive measure of influence in which connections from highly connected nodes confer greater importance than connections from peripheral ones^6^. Originally developed for web search, PageRank and related network-based methods have been used to recover prestige and hierarchy in citation networks, journals, academic genealogy, faculty hiring, and physician social networks^7–25^. Applied to residency placement, such methods offer a way to estimate how residency destinations are positioned within the observed system, rather than relying on external reputation measures alone.

This perspective may be especially useful in medical education, where institutional standing is widely invoked but difficult to define. Graduates of higher-ranked medical schools appear more likely to match into some competitive specialties^26,27^ or go on to have successful academic careers^28,29^, and residency leaders have suggested that school pedigree may play an even larger role in selection after the transition of USMLE Step 1 to pass/fail scoring^28,30^. At the same time, the rankings most often used as proxies for standing, particularly those from U.S. News & World Report (USNWR),^31^ remain highly influential despite longstanding criticism of their reliance on reputation surveys, editorial choices about which metrics to include, and arbitrary weighting schemes^32–34^. Alternative approaches have emphasized research productivity^34^, social mission^35^, or unsupervised clustering^36^, but they still depend on prior assumptions about which institutional attributes matter most. A network-based measure derived from actual match behavior offers a conceptually different approach: it estimates institutional position from observed placement flows.

Quantifying this network may also help address a separate but related question: how institutional advantage relates to specialty outcomes. Workforce projections continue to warn of major physician shortages, including in primary care^37^. At the same time, prior work has produced conflicting conclusions about whether educational debt pushes students away from lower-paying specialties and toward more highly remunerated fields^38–44^. This issue has drawn particular attention in discussions of tuition-free medical school and its potential effect on specialty choice^45–47^. A network-based model may help disentangle these relationships by testing whether specialty composition is associated with debt burden itself or instead reflects differences in access to influential residency pathways.

The major barrier to studying the Match in this way has been data availability. The NRMP reports aggregate outcomes but does not release the individual-level institutional pairings needed to reconstruct a national school-to-residency network. To address this limitation, we systematically aggregated publicly reported 2025 residency match outcomes from U.S. MD-granting medical schools and modeled the Match as a directed institutional network. We then asked whether network-derived measures recover meaningful dimensions of institutional influence and whether those measures help explain variation in specialty placement patterns across schools. To our knowledge, this is the first study to systematically analyze the U.S. residency match from a network perspective.

## METHODS

### Data collection and processing

The Association of American Medical Colleges (AAMC) reported 160 MD-granting programs in 2025^48,49^; five had not yet graduated a class, leaving 155 eligible. Institutions with multiple independently reporting campuses were treated as separate units, yielding 159 programs for analysis (eTable 1).

We aggregated 2025 residency match outcomes from publicly available school-published match lists, identified via targeted web searches. Of 155 eligible institutions, 114 provided accessible data in heterogeneous formats (HTML, PDF, image, or spreadsheet), parsed using an LLM-augmented pipeline (Gemini CLI 3.0, Google) into a standardized dataset with one row per matched student (home institution, residency destination, specialty). Manual review of a random subset (25 institutions, 3,412 records, 23.3% of the dataset) identified 23 mislabeled entries (error rate 0.68%), primarily from ambiguous or similarly named institutions (e.g., UT Health Sciences Houston vs San Antonio). Of the 114 institutions, 105 provided complete records (destination and specialty) and 9 provided partial records (destination only, n=2; specialty only, n=7); destination-only records were retained, and specialty-only records were excluded from network-based analyses. The resulting dataset comprised 107 institutions with usable match outcomes, broadly representative of all eligible programs across NIH funding, class size, indebtedness, and ranked specialties, with only modest differences in MCAT and GPA (eFigure 1).

### Network construction

The dataset was modeled as a directed weighted graph in which edges represent individual match outcomes from a medical school (source node) to a residency institution (target node), with edge weights corresponding to the number of students matching between each pair. After standardizing institution names, the final network included 107 medical schools and 301 residency institutions. Residency destinations were defined at the institutional level, with all programs within a given institution treated as a single node and affiliated institutions within the same health system consolidated.

### Network measures

Institutional position within the network was quantified using PageRank applied to the directed weighted graph, as implemented by the NetworkX^50^ python library. PageRank assigns higher scores to residency institutions that receive trainees from many institutions, particularly those that themselves recruit graduates from other highly connected institutions. In this setting, network position is inherently shaped by training capacity: programs with more positions recruit from a broader portion of the network and therefore occupy more central positions. Accordingly, PageRank reflects both the volume of trainees an institution receives and the distribution of their origins, assigning higher scores to programs that draw broadly from well-connected medical schools.

To evaluate medical school-level placement outcomes, we defined a placement score as the median PageRank percentile of residency destinations across a medical school’s graduating class. This measure summarizes the typical position of individual match outcomes within the network. Because it is defined at the level of individual placements and aggregated using the median, placement score is not determined by the size of residency programs, but instead reflects where students match within the observed distribution of training positions (eFigure 2).

For comparison, we evaluated alternative network centrality measures, including Katz centrality^51^, weighted and unweighted in-degree, weighted and unweighted out-degree, closeness centrality^52^, harmonic centrality^53^, betweenness centrality^54^, and HITS authority and hub scores^55^, all implemented in NetworkX^50^. These measures were computed on the same directed network and compared with external benchmarks.

### External validation

To assess whether network-derived measures captured meaningful dimensions of institutional position, institutional PageRank was compared with independent indicators, including 2025 NIH research funding^56^, the number of U.S. News & World Report–ranked specialties^57^, and Doximity residency reputation rankings^58^. For Doximity comparisons, analyses were restricted to institutions present in the rankings. Associations were quantified using Spearman correlation. School-level placement scores were similarly compared with academic selectivity metrics^59^, including mean MCAT score and undergraduate GPA.

### Stratified network analyses

For descriptive analyses of training pathways, medical schools and residency institutions were stratified into quintiles based on placement score and PageRank. Transition patterns were summarized by examining the distribution of matches across quintiles. To evaluate the contribution of home-program matches, we repeated analyses after excluding matches in which the residency institution corresponded to the student’s home medical school. Geographic movement was assessed by calculating straight-line (great-circle) distances between medical school and residency institution locations using latitude and longitude coordinates. Median relocation distance was summarized by specialty.

### Debt and specialty analyses

To evaluate the relationship between institutional debt burden and specialty composition, we examined associations between mean graduate indebtedness^60^ and the proportion of graduates entering primary care and surgical specialties. Associations were quantified using Spearman correlation. To assess whether these relationships were independent of institutional position within the match network, we performed adjusted analyses using linear regression models in which specialty proportions were modeled as a function of indebtedness with placement score included as a covariate. Residual associations between debt and specialty composition were then examined.

### Specialty definitions

Primary care specialties were defined as family medicine, internal medicine, pediatrics, and obstetrics–gynecology. Preliminary and transitional match years were included but analyzed as distinct categories, with preliminary medicine and preliminary surgery treated as separate fields. Specialty competitiveness was defined by unmatched rates among US MD seniors ranking that specialty as their only choice^5^; for non-NRMP specialties (e.g., urology, ophthalmology) and smaller unreported fields, competitiveness was approximated using the median unmatched rate among surgical or non-surgical specialties, as appropriate.

### Statistical analysis

Analyses were performed in Python (pandas, numpy, scipy, networkx, and statsmodels). Statistical tests were two-sided (α = 0.05); Spearman correlation coefficients are reported for nonparametric associations.

## RESULTS

We identified 114 U.S. MD-granting institutions with publicly available 2025 match data, of which 107 provided usable records, yielding 14,616 individual match outcomes (Figure 1). This corresponds to an estimated 73.5% of US MD seniors who matched nationally in 2025 (NRMP^5^, SF Match^61,62^, Urology match combined^63^, n = 19,878). Institutions with available data were broadly representative of all eligible programs, with only small differences in MCAT and GPA and no differences in indebtedness, NIH funding, class size, or number of ranked specialties (eFigure 1).

**Figure 1.**
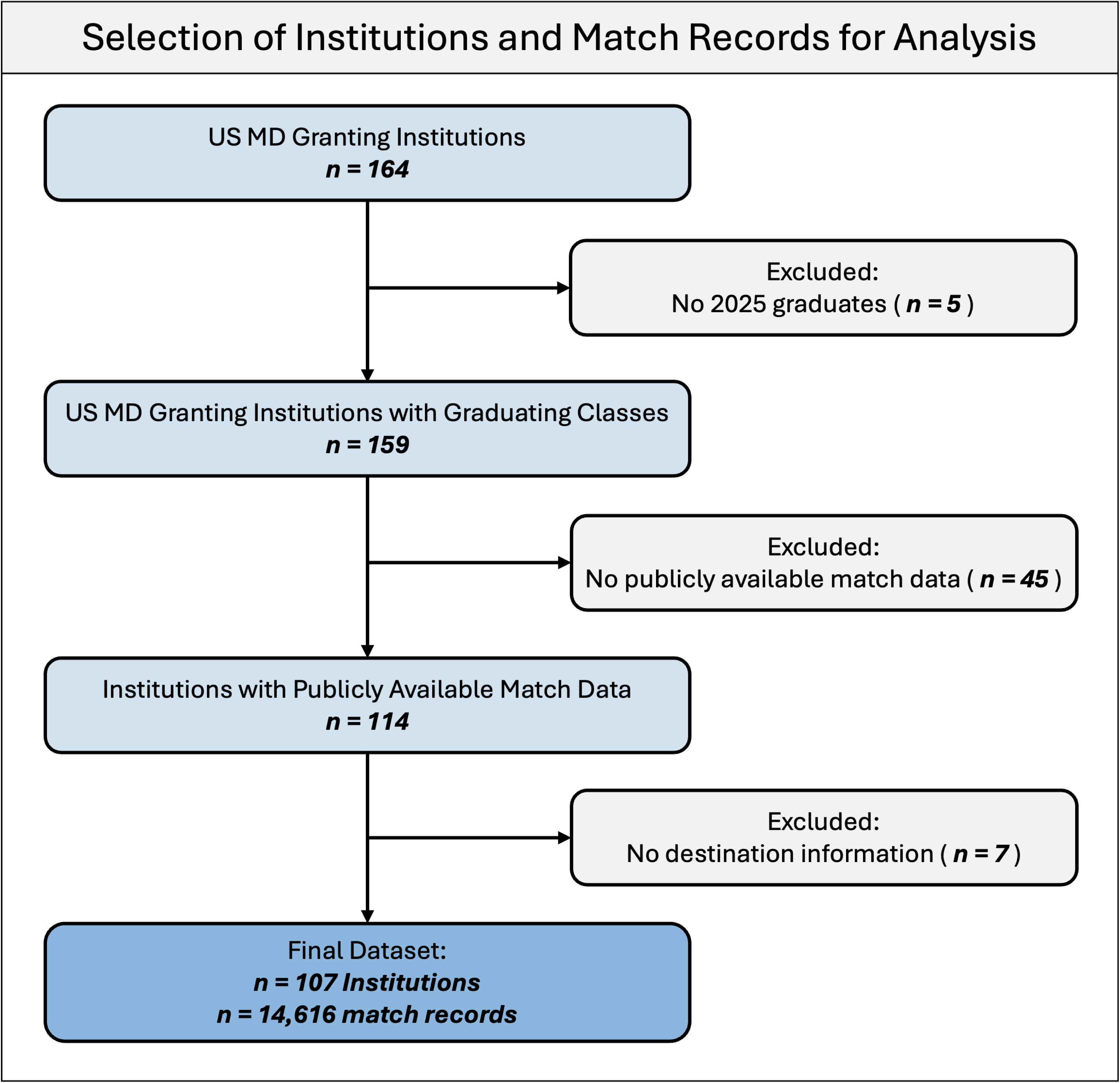
Identification and inclusion of 2025 residency match data. Flow diagram illustrating the identification, screening, and inclusion of publicly available match data from US MD-granting institutions. Of 159 total institutions with 2025 graduating classes, 114 provided publicly available match data, including 107 providing complete information on institutional destination. These data were aggregated to generate 14,616 individual match records.

Modeling the match as a directed network revealed a structured pattern of trainee movement between institutions (Figure 2A). We next evaluated network-derived centrality against independent benchmarks of institutional reputation and resources using Spearman rank correlation. PageRank-derived centrality of residency destinations was strongly associated with independent institutional characteristics, including NIH research funding (Spearman ρ = 0.85, p < 0.0001), Doximity residency reputation rankings (e.g., internal medicine, ρ = 0.86, p < 0.0001), number of ranked specialties by USNWR (ρ = 0.74, p < 0.0001) (Figure 2D). At the level of medical schools, we defined placement score as the median PageRank percentile of residency destinations across a school’s graduating class, capturing the extent to which students match into more central programs. Notably, while PageRank centrality of residency institutions reflects both connectivity and program size, the placement score captures the typical position of individual match outcomes and is therefore not inherently driven by institutional training capacity. This measure showed similar associations with institutional characteristics (MCAT ρ = 0.72, p < 0.0001; NIH funding ρ = 0.79, p < 0.0001). Concordance between PageRank and Doximity reputation rankings was observed across specialties (median ρ = 0.77), though the strength of association varied by field (eFigure 3), consistent with moderate inter-specialty agreement in reputation rankings (eFigure 4). Among alternative network measures, PageRank showed the most consistent associations across benchmarks (median ρ = 0.78; eFigure 5), supporting its use as a summary measure of institutional position within the match network.

**Figure 2.**
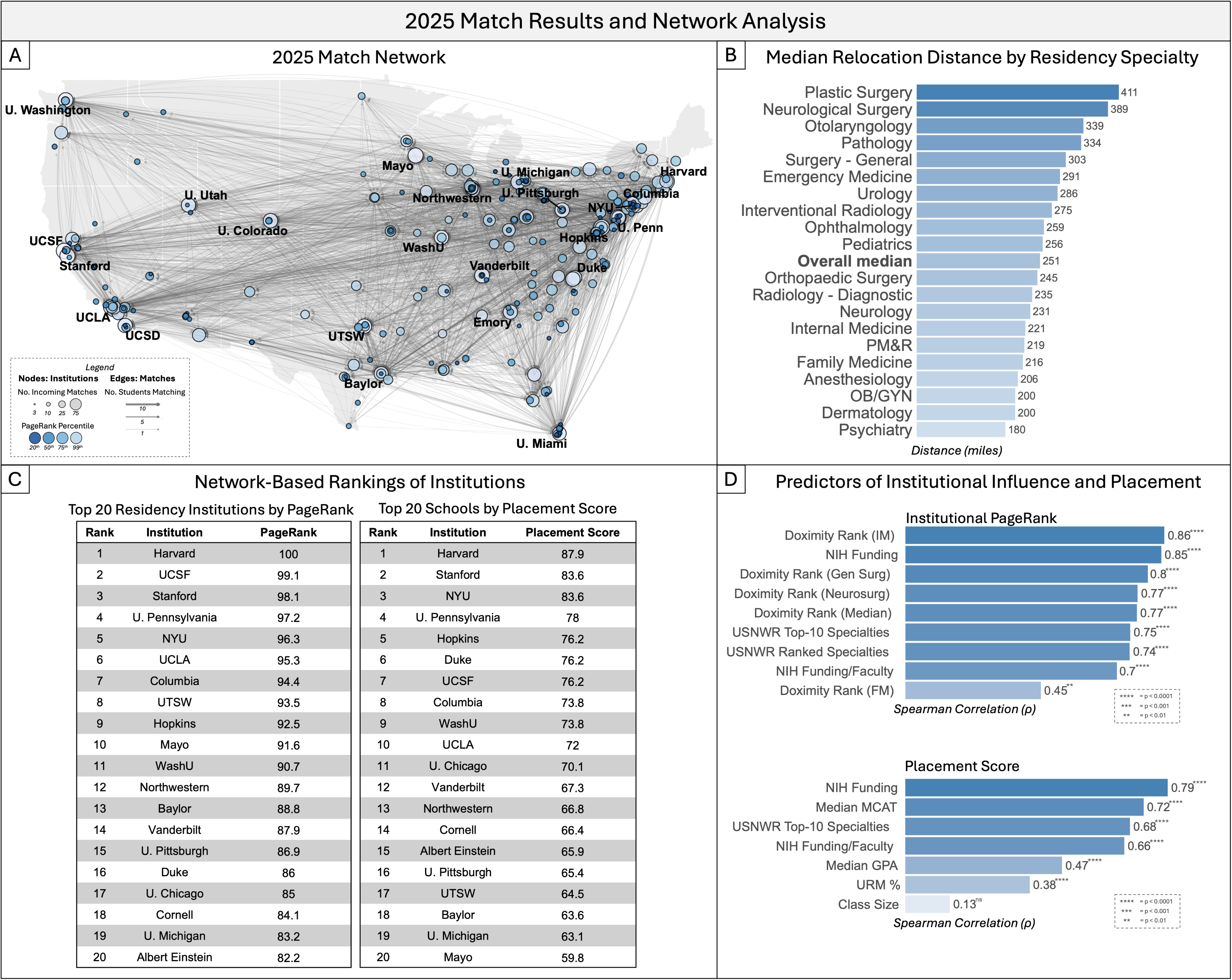
Network structure, geographic movement, and institutional predictors in the 2025 residency match. **(A)** Directed network of residency match flows between U.S. medical schools and residency institutions, with node size reflecting in-degree and color indicating PageRank-based institutional centrality. **(B)** Median relocation distance (miles) by specialty. **(C)** Network-based rankings of the top 20 residency institutions by PageRank percentile and medical school graduating class outcomes (placement score is defined as the median PageRank percentile of matched residency destinations). **(D)** Associations between network-derived measures and independent institutional characteristics, showing strong Spearman rank correlations with NIH research funding, survey-based residency reputation rankings (Doximity), U.S. News & World Report specialty rankings, and student academic metrics (e.g., MCAT, GPA).

Stratification by PageRank quintile revealed strong associations between medical school position and residency destination (Figure 3). Graduates of more central medical schools were substantially more likely to match into more central residency programs: 87.3% of students from top-quintile (Q1) schools matched into Q1 residencies compared to 41.0% from bottom-quintile (Q5) schools (Figure 3A). This pattern persisted after excluding home-program matches (82.8% vs 48.4%). Notably, many graduates from lower-ranked institutions still matched into highly connected programs. This partly reflects training-system structure: top-quintile schools sponsor disproportionately more residency positions, while lower-quintile schools may sponsor fewer programs and/or fill more slots via DO and international graduates; because our data cover only US MD graduates, we cannot distinguish between these possibilities. As a result, highly connected residency programs draw trainees from all quintiles, though disproportionately from more central institutions (Figure 3B; eFigure 6).

**Figure 3.**
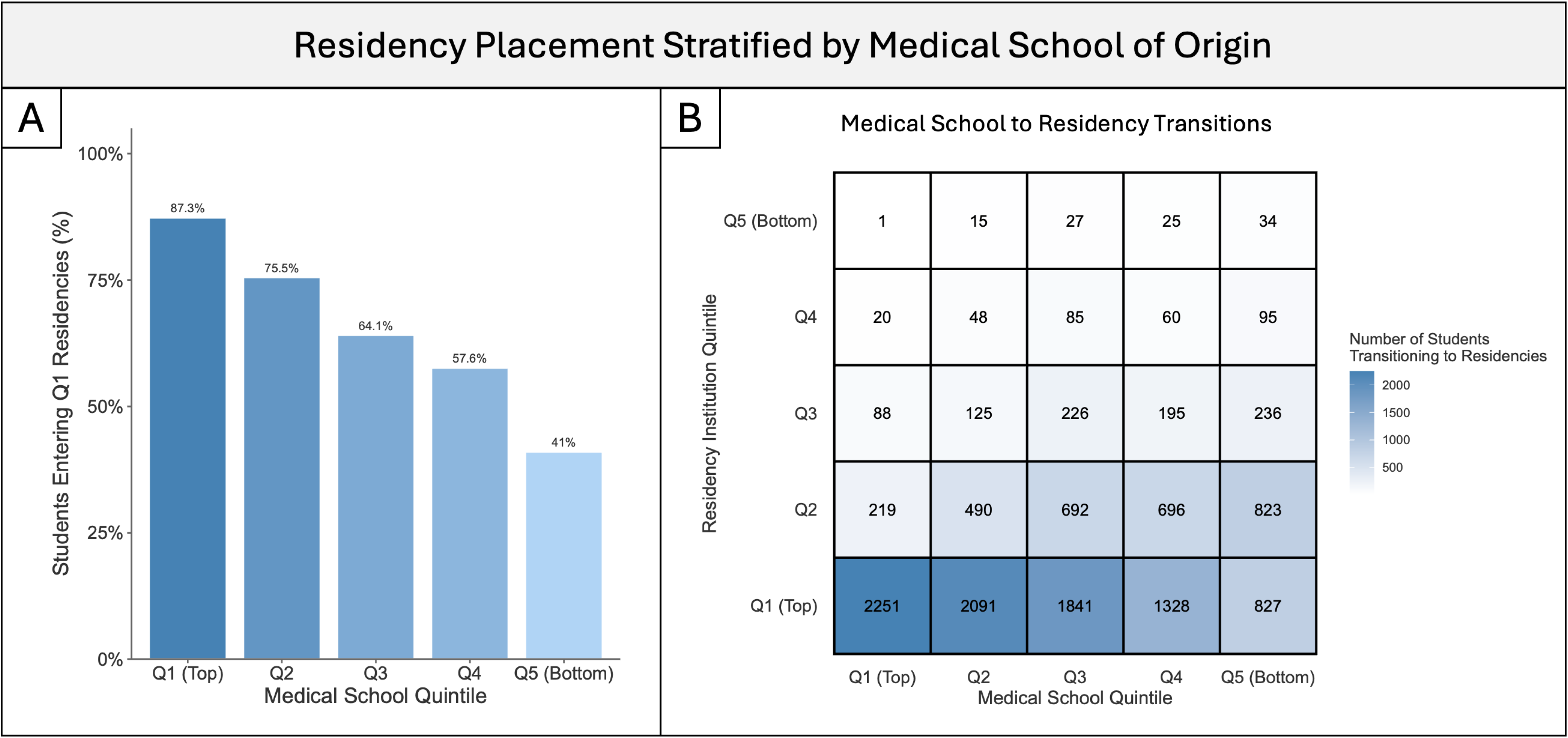
Residency placement stratified by institutional PageRank quintile. Medical institutions were stratified into PageRank quintiles. **(A)** Students from higher-ranked (Q1) medical schools were more likely to match into top-quintile (Q1) residency programs. **(B)** Transition matrix showing counts of MD students moving between quintiles. Q1 residency programs receive substantially more U.S. MD trainees within this dataset than Q5 programs. Because this network is constructed exclusively from available U.S. MD match outcomes, the observed lower volume of trainees entering Q5 programs reflects fewer placements from this specific cohort. This layout does not account for total institutional training capacity or the recruitment of osteopathic graduates and international medical graduates.

At the institutional level, placement score was associated with multiple graduate outcomes, including greater entry into surgical specialties (ρ = 0.60, p < 0.001), lower entry into primary care (ρ = −0.49, p < 0.001), higher average specialty competitiveness (ρ = 0.66, p < 0.001), and lower average graduate indebtedness (ρ = −0.36, p < 0.001) (eFigure 7). These associations were robust after excluding home-program matches (eFigure 8). Differences across schools were substantial: compared with bottom-decile institutions, top-decile institutions had higher proportions entering surgical specialties (29% vs 16%), lower proportions entering primary care (35% vs 47%), higher average specialty competitiveness (55th vs 46th percentile), and lower average indebtedness ($124K vs $197K) (Figure 4). Placement score captures a unifying dimension along which these differences organize, but should not be interpreted causally; whether they reflect institutional factors, student selection, or structural features of the training system cannot be determined.

**Figure 4.**
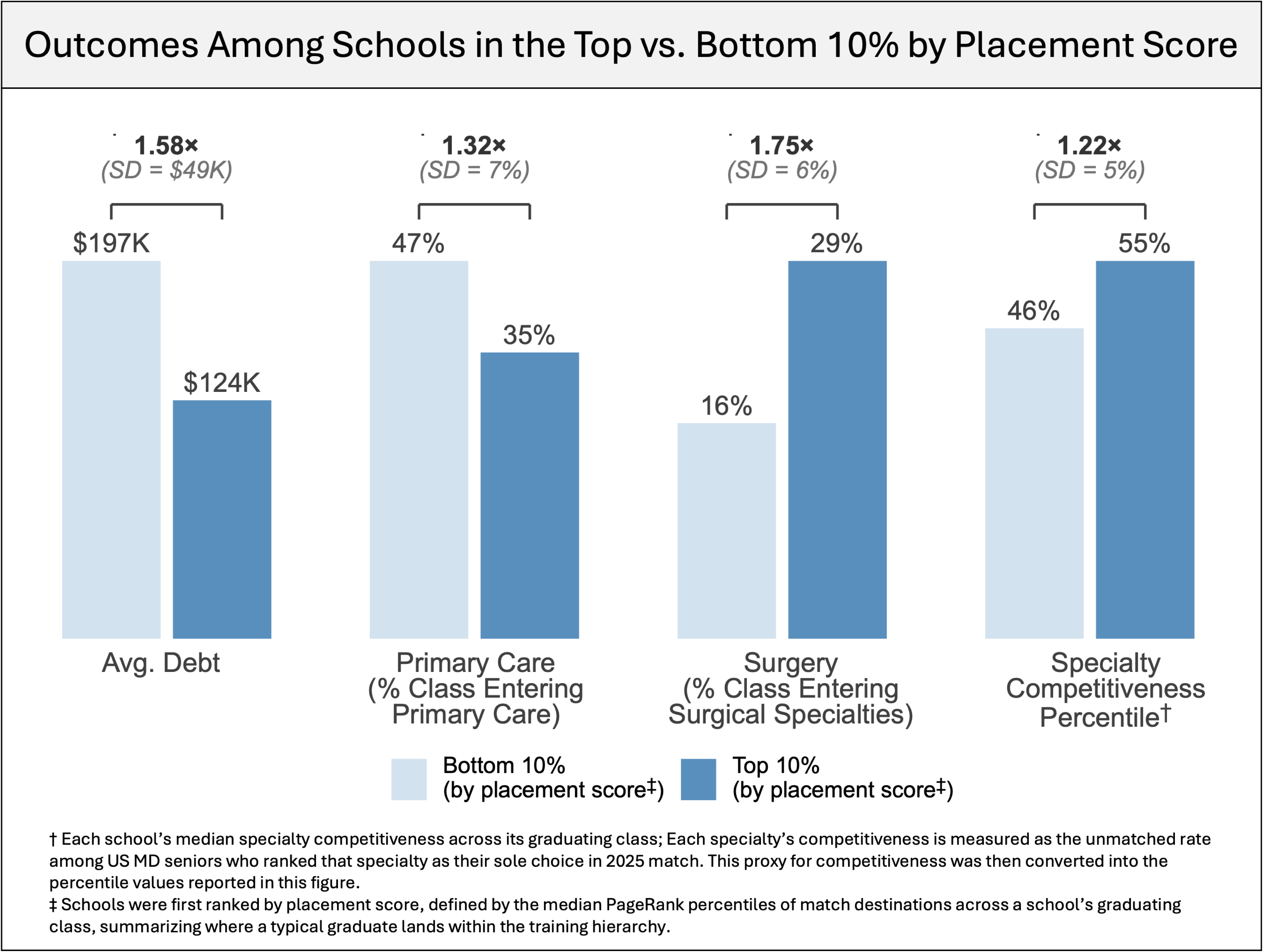
Variation in graduate outcomes between schools in the top and bottom decile by placement score. Medical schools were ranked by placement score (defined as the median PageRank percentile of match destinations across each school’s graduating class, summarizing where a typical graduate lands within the national training network) and outcomes were compared between schools in the top and bottom 10%. Schools in the top decile had substantially lower average graduate indebtedness ($124K vs. $197K), a lower proportion of graduates entering primary care (35% vs. 47%), a higher proportion entering surgical specialties (29% vs. 16%), and higher median specialty competitiveness (55th vs. 46th percentile). Ratios reflect the larger divided by the smaller mean; SD reflects the across-school standard deviation for each outcome among all 107 included institutions. Specialty competitiveness is measured as the median unmatched rate among U.S. MD seniors who ranked each specialty as their sole choice in the 2025 match, converted to a percentile across matched students.

Although prior work has proposed that higher educational debt may drive students away from primary care and toward higher-paying surgical specialties, evidence has been mixed^38–44^. In our data, unadjusted institutional-level analyses did not support this hypothesis: higher indebtedness was associated with greater entry into primary care (ρ = 0.20, p < 0.05) and lower entry into surgical specialties (ρ = −0.26, p < 0.01). No associations were observed after accounting for placement score, PageRank, and MCAT (eFigure 9). Interpretation is limited by strong intercorrelations among institutional characteristics, which complicate isolation of an independent debt effect; adjustment may capture shared upstream influences while attenuating potential pathways (eFigure 10). Overall, these findings do not support a positive association between educational debt and entry into higher-paying specialties at the institutional level.

## DISCUSSION

We reconstructed the 2025 US residency match for MD graduates as a national institutional network using publicly reported outcomes, capturing an estimated 73.5% of the 2025 national match cohort. Network-derived measures of institutional position aligned closely with independent indicators of standing, including NIH funding, specialty rankings, residency reputation, and student selectivity. These findings suggest that patterns of trainee movement within the residency match capture a coherent gradient of institutional position.

Consistent with this structure, graduates of higher-positioned medical schools experienced systematically different outcomes than those of lower-positioned schools. Although such differences are often assumed, they have been difficult to characterize quantitatively at a national scale. Compared with bottom-decile institutions, top-decile institutions had higher rates of surgical specialty entry, lower rates of primary care entry, higher specialty competitiveness, and lower graduate indebtedness (Figure 4). These differences may reflect institutional factors, student selection, or structural features of the training system, which cannot be disentangled in these data. Similar interpretive challenges have been observed in higher education, where apparent advantages of selective institutions may reflect differences in admitted students rather than institutional effects^64,65^. Disentangling these contributions would require individual-level data on applicant characteristics, preferences, and outcomes.

These results have implications for how institutional position is conceptualized and measured. Although existing ranking systems are widely used, they rely heavily on reputation surveys and subjective weighting^33,34^. While our analysis does not establish that network-derived measures are preferable, it shows that placement patterns alone recover much of the same structure. Because these measures arise from observed institutional relationships, they provide a complementary, behavior-based perspective on institutional position. Importantly, PageRank centrality reflects both connectivity and training capacity: programs with more positions recruit more broadly and occupy more central positions. Capacity is therefore not a confounder but a defining feature of the system, and our findings should be interpreted as describing how training opportunities are structured and distributed, rather than isolating prestige independent of program size.

We also observed strong convergence across domains often considered distinct. Research funding, clinical reputation, student selectivity, and residency placement outcomes were highly correlated, suggesting these characteristics tend to co-occur in practice. The close association with NIH funding is notable, since medical student training is not primarily research-focused.

At the institutional level, we found no evidence that higher educational debt drives students toward higher-paying specialties. Instead, variation in aggregate graduate outcomes was not independently associated with debt after accounting for institutional position. This does not exclude a role for debt in individual decision-making, but suggests that school-level differences in specialty composition are not primarily driven by financial burden.

## LIMITATIONS

Several limitations should be considered. First, our analysis was restricted to US MD graduates and excluded DO and international medical graduates (IMGs), who together fill a substantial share of US residency positions; institutional patterns among these groups may differ from those observed here and warrant separate study. Second, analyses were conducted at the institutional level and do not account for individual applicant characteristics; because institutional position correlates with student selectivity, observed associations likely reflect both institutional context and underlying differences in trainees. Third, the analysis relied on publicly available match lists, which may introduce bias if institutions with more favorable outcomes were more likely to report data. Fourth, the study reflects a single match cycle and provides a cross-sectional view of a dynamic system. Fifth, residency destinations were analyzed at the institutional rather than program level, improving interpretability but reducing granularity. Finally, placement score and PageRank are modeled summary measures of position within observed placement patterns and should not be interpreted as definitive assessments of institutional performance, training quality, or causal influence.

Despite these limitations, this study demonstrates that publicly available match outcomes can characterize the residency system at a national scale. With longitudinal data, future work could assess the stability of institutional position over time and evaluate how changes in training pathways relate to specialty choice and workforce outcomes.

## CONCLUSIONS

In this analysis of the 2025 US residency match, capturing an estimated 73.5% of the national MD match cohort, institutional position was closely tied to residency placement, educational debt, and specialty choice, reflecting a hierarchical structure in which graduates of higher- and lower-positioned medical schools experience systematically different outcomes. These findings offer a population-level, behavior-based perspective on how institutional standing shapes training pathways.

## Supporting information

eMaterials

## Data Availability

All data analyzed and presented herein were publicly available prior to publication. NIH funding data were obtained from the Blue Ridge Institute for Medical Research. Medical school characteristics, including MCAT scores, GPA, and graduate indebtedness, were obtained from the AAMC Medical School Admission Requirements database. Institutional listings and matriculant demographics were obtained from AAMC reports. U.S. News & World Report 2025 hospital and specialty rankings were used for institutional comparisons, and residency reputation rankings were obtained from Doximity. Publicly available residency match data sources for each institution were collected online as of March 2025; these sources including links to public URLs are detailed in eTable 1. Code and processed data are available from the authors upon reasonable request.

https://brimr.org/brimr-rankings-of-nih-funding-in-2025/

https://mec.aamc.org/msar-ui/

https://www.aamc.org/data-reports/students-residents/data/facts-applicants-and-matriculants

https://www.aamc.org/data-reports/students-residents/data/facts-enrollment-graduates-and-md-phd

https://www.usnews.com/info/blogs/press-room/articles/2025-07-29/u-s-news-announces-2025-2026-best-hospitals

https://www.doximity.com/residency/

https://www.nrmp.org/match-data/2025/05/results-and-data-2025-main-residency-match/

## Conflict of Interest Disclosures

Dr Burk-Rafel reported consulting for ScholarRx and Liaison International. All other authors report no conflicts of interest.

## Funding/Support

None reported

## Author Contributions

JIT has full access to all of the data in the study and takes responsibility for the integrity of the data and the accuracy of the data analysis.

## Concept and design

JIT

## Acquisition, analysis, or interpretation of data

JIT, AA, JBR, EKO

## Drafting of the manuscript

JIT, with feedback from all authors

## Critical review of the manuscript for important intellectual content

JIT, AA, JBR, EKO

## Use of Artificial Intelligence

Gemini (version 3.0; Google) was used to assist with parsing and standardizing publicly available match list data from heterogeneous formats into a structured dataset, as described in the Methods, and for limited language editing. All study design, data review, statistical analysis, interpretation of findings, literature review, figure construction, and reference selection were performed exclusively by the human authors. All AI-assisted outputs were reviewed and fully validated by the authors, who take full responsibility for the accuracy and integrity of the manuscript.

## Data Sharing Statement

All data analyzed and presented herein were publicly available prior to publication. NIH funding data were obtained from the Blue Ridge Institute for Medical Research^56^. Medical school characteristics, including MCAT scores, GPA, and graduate indebtedness, were obtained from the AAMC Medical School Admission Requirements database^59,60^. Institutional listings and matriculant demographics were obtained from AAMC reports^48,49^. U.S. News & World Report 2025 hospital and specialty rankings were used for institutional comparisons^57^, and residency reputation rankings were obtained from Doximity^58^. Publicly available residency match data sources for each institution were collected online as of March 2025; these sources including links to public URLs are detailed in eTable 1. Code and processed data are available from the authors upon reasonable request.

## Notes

### Competing Interest Statement

Dr Burk-Rafel reported consulting for ScholarRx and Liaison International. Mr. Turner, Mr. Arias and Dr. Oermann report no conflicts of interest.

### Author Declarations

The study used ONLY openly available data that were originally located from a number of sources already available on the internet. This study did not involve identifiable human subject data, clinical or otherwise. We pooled de-identified information about graduating medical students' residency match destinations from official, publicly available sources as of March 2025; these sources, including links to public URLs, are detailed in eTable 1. See Data Availability statement for more details.

