## Supplementary material for "Institutional Standing and Trainee Outcomes in the 2025 US Residency Match": eMaterials

### **eTable 1****. U.S. MD-Granting Medical Schools Included in the Institutional Match Network Analysis, and Links to Publicly Available 2025 Match Data**

### **eFigure 1****. Comparison of institutional characteristics between programs with and without publicly available match data**

### **eFigure 2****. Schematic Overview of PageRank**

### **eFigure 3****. Correlation of PageRank with Doximity residency specialty rankings**

### **eFigure 4****. Spearman correlation matrix of Doximity Rankings across specialties**

### **eFigure 5****. Performance of alternative network centrality measures**

### **eFigure 6****. Origin of incoming PGY1 residents at top-quintile residency programs**

### **eFigure 7****. Institutional placement score predicts debt and specialty composition**

### **eFigure 8****. Placement score and specialty composition after excluding home-program matches**

### **eFigure 9****. Association of educational debt with specialty composition before and after accounting for network position**

### **eFigure 10****. Proposed causal framework linking institutional resources, graduate indebtedness, and specialty outcomes**

### **eTable 1. U.S. MD-Granting Medical Schools Included in the Institutional Match Network Analysis, and Links to Publicly Available 2025 Match Data**

Publicly available links to institutional Match Day results or match lists are provided where identified as of March 2026; a dash indicates that no publicly available link could be identified. N = 159 programs.

| **Institution** | **URL** |
| --- | --- |
| Alabama-Heersink | <https://www.uab.edu/medicine/home/images/match-results-by-student-2025.pdf> |
| Albany | <https://www.albanymed.org/wp-content/uploads/sites/2/2025/03/2025_residency-positions-booklet_3col_v3.pdf> |
| Arizona | *—* |
| Arizona Phoenix | <https://www.facebook.com/UAZMEDPHX/posts/nearly-two-weeks-ago-matchday2025-occurred-and-it-was-an-incredible-success-for-/1290669755958661/> |
| Arkansas | <https://medicine.uams.edu/students/preparing-for-residency/residency-match-2025/> |
| Baylor | <https://blogs.bcm.edu/2025/03/21/celebrating-match-day-2025/> |
| Brown-Alpert | <https://medical.brown.edu/md-2025-match-list> |
| BU-Chobanian Avedisian | <https://www.bumc.bu.edu/camed/files/2025/04/Match-numbers-in-each-specialty-requested-numbers-withheld.pdf> |
| Buffalo-Jacobs | <https://medicine.buffalo.edu/news_and_events/news/2025/3/match-day-2025-22088.html> |
| California | <https://cusm.edu/academics/md/_archive/md-students/Match%20Data%202025.pdf> |
| California Northstate | <https://medicine.cnsu.edu/PDFs/2025_Match_Results.pdf> |
| Caribe | *—* |
| Carle Illinois | <https://medicine.illinois.edu/news/residency-match-results-ci-med-class-of-2025-lands-prestigious-programs-and-specialties> |
| Case Western Reserve | <https://case.edu/medicine/md/sites/default/files/2025-07/CWRU%20Match%20List%202025%20-%20De-Identified%20%283%29.pdf> |
| Case Western Reserve Lerner | <https://portal.cclcm.ccf.org/cclcm/CCLCMDependencies/popup/match_2025.html> |
| Central Michigan | *—* |
| Chicago Med Franklin | <https://rfums-bigtree.s3.amazonaws.com/files/resources/cms-match-list-2025.pdf> |
| Chicago-Pritzker | <https://d54gi6idwcev6.cloudfront.net/sites/pritzker/files/2025-03/Anonymized%20Match%20List%20for%20Web%20-%2025.pdf> |
| Cincinnati | <https://comdo-wcnlb.uc.edu/emos/studentservices/MatchSummary.aspx> |
| Colorado | <https://medschool.cuanschutz.edu/docs/librariesprovider31/education-docs/match-day/full-public-match-day-list---3-20-25-5.pdf?sfvrsn=fce123b4_1> |
| Columbia-Vagelos | <https://www.vagelos.columbia.edu/education/student-resources/office-student-affairs/match-day> |
| Connecticut | *—* |
| Cooper Rowan | <https://cmsru.rowan.edu/documents/alumni-documents/residencies/residency-listing-2025.pdf> |
| Cornell-Weill | <https://medicaleducation.weill.cornell.edu/sites/default/files/class2025_match_results.pdf> |
| Creighton | <https://www.creighton.edu/sites/default/files/match-list-2025.pdf> |
| CUNY | *—* |
| Dartmouth-Geisel | <https://geiselmed.dartmouth.edu/admissions/wp-content/uploads/sites/3/2025/04/2025-Internship-Residency-Results-by-Program.pdf> |
| Drexel | <https://drexel.edu/medicine/academics/md-program/residency-match/> |
| Duke | <https://medschool.duke.edu/news/duke-school-medicine-match-day-2025> |
| East Carolina-Brody | <https://medicine.ecu.edu/data-analysis-strategy/residency-program-match-results/> |
| East Tennessee-Quillen | <https://www.etsu.edu/com/admissions/collegeinfo/placement-results-25.php> |
| Eastern Virginia ODU | <https://www.evms.edu/media/Match_list_2025_FINAL.pdf> |
| Einstein | <https://einsteinmed.edu/education/md-program/md-admissions/md-admissions-statistics/match-day-results> |
| Emory | *—* |
| FIU-Wertheim | <https://medicine.fiu.edu/resources/current-students/md-resources/md-events/_assets/docs/match-day-2025-results.pdf> |
| Florida | <https://osa.med.ufl.edu/wordpress/files/2025/05/Co2025-Match-Results-for-Website.pdf> |
| Florida Atlantic-Schmidt | *—* |
| Florida State | <https://med.fsu.edu/sites/default/files/userFiles/2025%20Match%20Day%20Results.pdf> |
| Geisinger Commonwealth | *—* |
| George Washington | <https://public.tableau.com/app/profile/gw.business.intelligence.services/viz/SMHSMatchDay2025/MappedMatchDayMapofStudents> |
| Georgetown | <https://meded.georgetown.edu/admissions/degrees-and-admissions/md/matchplacement/> |
| Hackensack Meridian | <https://www.hmsom.edu/en/admissions/match-data> |
| Harvard | <https://hms.harvard.edu/sites/default/files/2025-10/match_list_de-identified_2025_for_website.pdf> |
| Hawaii-Burns | *—* |
| Houston-Fertitta | <https://stories.uh.edu/2026-match-day/assets/ViA0bTrMlR/2025-public-match-results.pdf> |
| Howard | *—* |
| Illinois | <https://chicago.medicine.uic.edu/depts/support-offices/osa/match-results/> |
| Illinois | <https://peoria.medicine.uic.edu/campus/student-life/ceremonies-and-events/match-day-results/> |
| Illinois | <https://rockford.medicine.uic.edu/depts/offices/office-of-student-affairs/match-day/> |
| Indiana | *—* |
| Iowa-Carver | <https://md.medicine.uiowa.edu/sites/md.medicine.uiowa.edu/files/2025-05/2025_MatchTable6_Specialty_pub.pdf> |
| Jefferson-Kimmel | <https://www.jefferson.edu/content/dam/academic/skmc/admissions/skmc-admissions-interview-day-match-list-0525.pdf> |
| Johns Hopkins | <https://www.scribd.com/document/862129221/2025-Match-List-Public> |
| Kaiser Permanente-Tyson | <https://medschool.kp.org/about/match-results> |
| Kansas | *—* |
| Kentucky | <https://www.lexingtondoctors.org/wp-content/uploads/2025/03/Match-list-2025.pdf> |
| UNLV-Kerkorian | <https://www.unlv.edu/medicine/md-program/match-day/2025> |
| Loma Linda | *—* |
| Louisville | <https://louisville.edu/medicine/admissions/files/2025-match-results/> |
| Loyola-Stritch | *—* |
| LSU New Orleans | <https://www.medschool.lsuhsc.edu/student_affairs/match.aspx> |
| LSU Shreveport | <https://schoolofmedicine.lsuhs.edu/current-students/events/match-day/2025-match-day> |
| Marshall-Edwards | <https://jcesom.marshall.edu/media/63727/2025-jcesom-residency-match-list.pdf> |
| Maryland | *—* |
| Massachusetts-Chan | *—* |
| Mayo | <https://college.mayo.edu/media/mccms/content-assets/about/commencement/2025-Mayo-Clinic-Commencement-Program-MC1603rev042925.pdf> |
| MC Georgia Augusta | <https://augustauniversity.app.box.com/s/2fo7plagnyh0hyq3raoucjvbdp22eg2r> |
| MC Wisconsin | <https://secondaryapplication.mcw.edu/JQFileUpload.ashx?_method=GET&folder=&file=MCW+Match+Results+2025.pdf> |
| Meharry | *—* |
| Mercer | *—* |
| Miami-Miller | <https://med.miami.edu/medical-education/student-affairs/match-day-results> |
| Michigan | <https://medschool.umich.edu/match-list?field_year_match_value=All&field_name_pgy2_match_value=&field_specialty_match_target_id=All> |
| Michigan State | *—* |
| Minnesota | <https://med.umn.edu/sites/med.umn.edu/files/2025-03/Class%202025%20Match%20List%20and%20Statistics.pdf> |
| Mississippi | <https://umc.edu/som/Students/Career%20Advising%20Resources/Results/2020s/2025-Residency-Match-Results.html> |
| Missouri Columbia | <https://medicine.missouri.edu/offices-programs/admissions/match-lists> |
| Missouri Kansas City | <https://med.umkc.edu/news-events/featured-events-lectures/match-day/match-2025-kansas-city.html> |
| Missouri Kansas City | <https://med.umkc.edu/news-events/featured-events-lectures/match-day/match-2026-st-joseph.html> |
| Morehouse | *—* |
| Mount Sinai-Icahn | *—* |
| MU South Carolina | *—* |
| Nebraska | <https://pdfhost.io/v/LxN6ygpbZH_Nebraska_Match_2025> |
| Nevada Reno | *—* |
| New Mexico | *—* |
| New York Medical | <https://www.nymc.edu/som/about/outcomes/2025-match-day/> |
| North Carolina | <https://www.med.unc.edu/md/events/wp-content/uploads/sites/1378/2025/04/2025-Match-by-Specialty.pdf> |
| North Dakota | <https://med.und.edu/admissions/student-affairs-admissions/_files/match-lists/match-list-2025.pdf> |
| Northeast Ohio | <https://www.neomed.edu/medicine/match-day-25/> |
| Northwestern-Feinberg | <https://www.feinberg.northwestern.edu/md-education/current-students/career-development/residency-application-process/match-results/index.html> |
| Nova Southeastern-Patel | *—* |
| NYU Long Island-Grossman | <https://web.archive.org/web/20250810100755/https://medli.nyu.edu/education/md-degree/md-admissions/match-day-results> |
| NYU-Grossman | <https://med.nyu.edu/education/md-degree/md-admissions/match-day-results> |
| Oakland Beaumont | <https://www.oakland.edu/Assets/Oakland/medicine/graphics/Match-Day/032125%20MatchDay2025.pdf> |
| Ohio State | *—* |
| Oklahoma | <https://medicine.ouhsc.edu/Portals/1365/Assets/OKC%20Public%20Match%20List%202025.pdf> |
| Oregon | <https://s3.amazonaws.com/cms.ipressroom.com/296/files/20252/2025+OHSU+Match+Day+Stats.pdf> |
| Penn State | <https://med.psu.edu/education/degree-programs/md-doctor-medicine/residency-matches> |
| Pennsylvania-Perelman | <https://www.med.upenn.edu/admissions/assets/user-content/documents/2025-graduating-residency-outcomes.pdf> |
| Pittsburgh | <https://www.medadmissions.pitt.edu/sites/default/files/assets/2025%20MATCH%20LIST%20-%20INSTITUTIONS%20ONLY.pdf> |
| Ponce | <https://phsu.edu/_resources/documents/Match%20Data%202025-PHSU.pdf> |
| Puerto Rico | *—* |
| Quinnipiac-Netter | <https://medicine.qu.edu/programs/medical-doctor-degree/md/residency-match/2025-match-day/> |
| Renaissance Stony Brook | <https://pdfhost.io/v/M6HuuChpSK_StonyBrook2025Match> |
| Rochester | *—* |
| Rush | *—* |
| Rutgers New Jersey | <https://njms.rutgers.edu/admissions/documents/2025ResidencyPlacementbySpecialty.pdf> |
| Rutgers-RW Johnson | <https://rwjms.rutgers.edu/sites/default/files/2025-03/Match%20list%202025_Public-final.pdf> |
| Saint Louis | *—* |
| San Juan Bautista | *—* |
| South Alabama-Whiddon | <https://www.southalabama.edu/colleges/com/currentstudents/resources/match-day-results-2025.pdf> |
| South Carolina Columbia | <https://www.sc.edu/study/colleges_schools/medicine/documents/2025_match_results.pdf> |
| South Carolina Greenville | <https://sc.edu/study/colleges_schools/medicine_greenville/docs/match_day_specialities_2025.pdf> |
| South Dakota-Sanford | <https://www.usd.edu/academics/colleges-and-schools/sanford-school-of-medicine/south-dakotan-medicine/usd-medical-students-learn-post-graduate-destinations-at-match-day-2025> |
| Southern Cal-Keck | *—* |
| Southern Illinois | <https://www.siumed.edu/news/siu-school-medicine-residency-match-results-class-2025> |
| Stanford | <http://med.stanford.edu/content/dam/sm-news/content/2025/04/residency-placement-outcomes-2025.pdf> |
| SUNY Downstate | <https://www.downstate.edu/education-training/student-affairs/residency-placement-lists/2025.html> |
| SUNY Upstate-Norton | <https://www.upstate.edu/com/pdf/2025-match-list.pdf> |
| TCU-Burnett | <https://mdschool.tcu.edu/alumni/match-map/> |
| Temple-Katz | <https://medicine.temple.edu/sites/medicine/files/media/document/rpt_byinstitution_forwebsite_20250502.pdf> |
| Tennessee | *—* |
| Texas A&M | <https://pdfhost.io/v/z7W9yb7eju_TAMU_2025_Match_List__2_> |
| Texas Tech | <https://www.ttuhsc.edu/medicine/student-affairs/documents/MatchListClassof2025-Public_03212025.pdf> |
| Texas Tech-Foster | <https://ttuhscep.edu/som/studentaffairs/match-day-results/_documents/2025_Match_List_FINAL.pdf> |
| Toledo | <https://www.utoledo.edu/med/md/pdfs/utoledo-match-2025-summary.pdf> |
| Tufts | <https://medicine.tufts.edu/academics/medicine/match-results> |
| Tulane | <https://medicine.tulane.edu/sites/default/files/2025-04/FINAL--2025-Match-List-Results.pdf> |
| U Washington | <https://education.uwmedicine.org/career-advising/wp-content/uploads/sites/4/2025/04/2025-Match-Results_For-Public-Distribution.pdf> |
| UC Davis | <https://health.ucdavis.edu/mdprogram/registrar/RegistrarForms/match-results.pdf> |
| UC Irvine | *—* |
| UC Riverside | *—* |
| UC San Diego | *—* |
| UC San Francisco | <https://meded.ucsf.edu/about-us/program-statistics/residency-match-outcomes> |
| UCF | <https://med.ucf.edu/media/2025/03/Match-Results-2025-BY-SPECIALTY.pdf> |
| UCLA-Geffen | <https://medschool.ucla.edu/sites/g/files/oketem456/files/media/documents/2025_match_list_-_for_website.pdf> |
| Uniformed Services-Hebert | *—* |
| USF-Morsani | <https://online.pubhtml5.com/vclzp/ybdp/> |
| UT Austin-Dell | <https://dellmed.utexas.edu/education/academics/undergraduate-medical-education/residency-match-results> |
| UT Houston-McGovern | <https://drive.google.com/file/d/1A86tgg-40OTAOiwR91JAX4_-fK5SkQN_/view> |
| UT Medical Branch-Sealy | <https://www.utmb.edu/som/student-affairs/events/match-day-ceremony> |
| UT Rio Grande Valley | <https://www.utrgv.edu/som/admissions/match-day/2025/index.htm> |
| UT San Antonio-Long | <https://uthscsa.edu/medicine/education/ume/admissions/residency-match-results> |
| UT Southwestern | <https://www.utsouthwestern.edu/ctplus/stories/2025/match-day-2025-list.html> |
| Utah-Eccles | <https://medicine.utah.edu/documents/uusom-2025-match-report> |
| Vanderbilt | <https://cdn.vanderbilt.edu/t2-main/medschool-prd/wp-content/uploads/sites/49/2025/03/VUSM_2025_Match-Results_releasable.pdf> |
| Vermont-Larner | <https://www.uvm.edu/d10-files/documents/2025-03/Public-Match-List-co2025.pdf> |
| Virginia | <https://med.virginia.edu/md-program/student-affairs/student-resources/residency-match-results/> |
| Virginia Commonwealth | <https://medschool.vcu.edu/education/medical-education/match-day/2025/match-list/> |
| Virginia Tech Carilion | *—* |
| Wake Forest | <https://cdn.atriumhealth.org/-/media/wakeforest/school/files/education-and-training/md-program/match-day/match-day-2025-match-locations.pdf?rev=d40e494b6acc4a41b88e7830c9b1ce0d> |
| Washington State-Floyd | *—* |
| Washington U St Louis | <https://mdadmissions.wustl.edu/education/student-outcomes/> |
| Wayne State | <https://www.med.wayne.edu/match-day/pdf/wsu-som-public-match-list-2025.pdf> |
| West Virginia | <https://medicine.wvu.edu/md-student-services/match-day/2025/> |
| Western Michigan-Stryker | <https://wmed.edu/sites/default/files/Class%20of%202025%20Match%20Results.pdf> |
| Wisconsin | *—* |
| Wright State-Boonshoft | <https://medicine.wright.edu/alumni-and-giving/residency-match-results> |
| Yale | <https://medicine.yale.edu/news-article/yale-school-of-medicine-celebrates-exemplary-matches/> |
| Zucker Hofstra Northwell | <https://medicine.hofstra.edu/sites/medicine.hofstra.edu/files/2025-03/matchday-graphic-2025.png> |

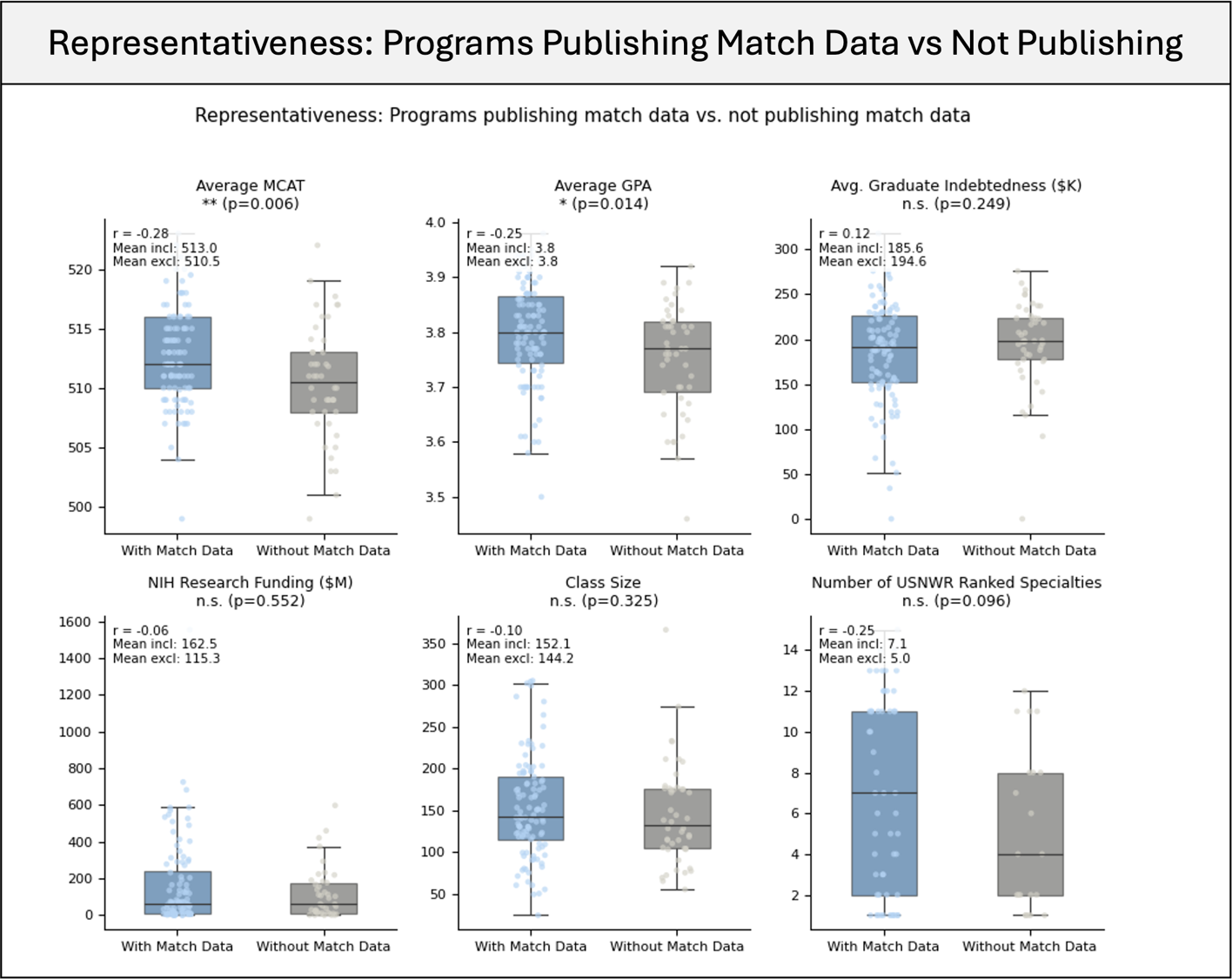

**eFigure 1. Comparison of institutional characteristics between programs with and without publicly available match data.** Each point represents a U.S. MD-granting institution. Box plots show the median and interquartile range; overlaid points represent individual institutions (jittered for visibility). Groups were compared using two-sided Mann-Whitney U tests, with rank-biserial effect sizes and p values plotted. Programs with publicly available match data differed significantly from those without on Average MCAT (median 513.0 vs. 510.5; p=0.006) and Average GPA (median 3.80 vs. 3.77; p=0.014), with institutions publishing match data scoring modestly higher on both metrics; however, effect sizes for both comparisons were small (rank-biserial r = −0.28 and −0.25, respectively). No significant differences were observed for average graduate indebtedness, NIH research funding, graduating class size, or number of U.S. News & World Report–ranked specialties.

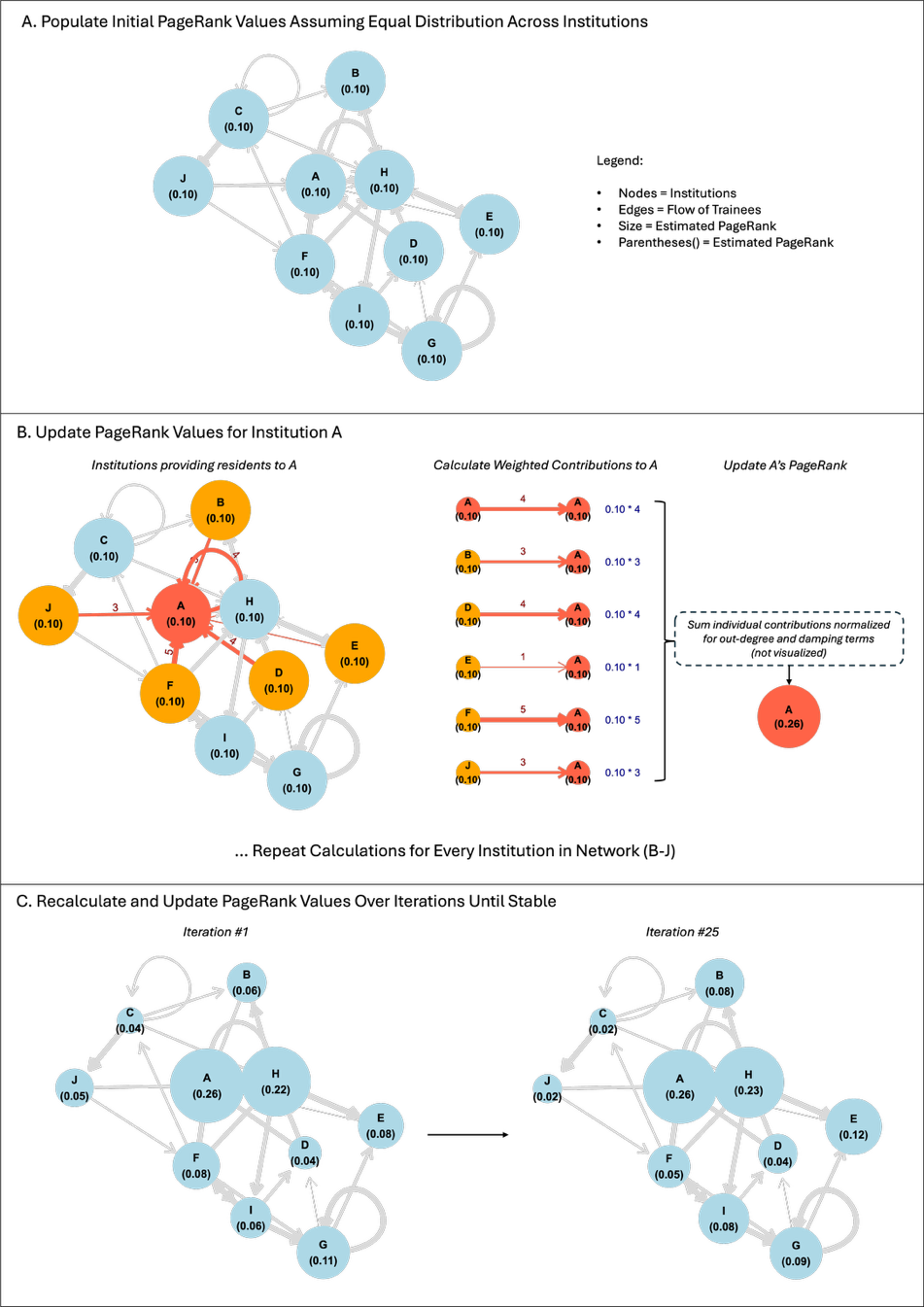

**eFigure 2: Schematic Overview of PageRank
(A)** The residency match is modeled as a directed, weighted graph where nodes represent institutions and edge weights represent trainee volumes. PageRank values are initially distributed uniformly across all N institutions (1 / N = 0.10 for this 10-node toy network)
(**B)** The updated PageRank for a single node (e.g., Institution A) is computed by aggregating incoming trainee flows from its sending institutions. Each sender's contribution is scaled by the specific edge weight and normalized by its total out-degree. This model incorporates auto-connections (self-loops), allowing home-program retention to contribute directly to an institution's score. The resulting sum is adjusted by a standard damping factor (0.85) to distribute a baseline probability mass uniformly across the network.
(**C**) The node-level calculations are repeated simultaneously for all institutions across successive iterations. Values redistribute based on network topology until they reach a steady state, yielding the final institutional PageRank scores used to quantify network centrality.

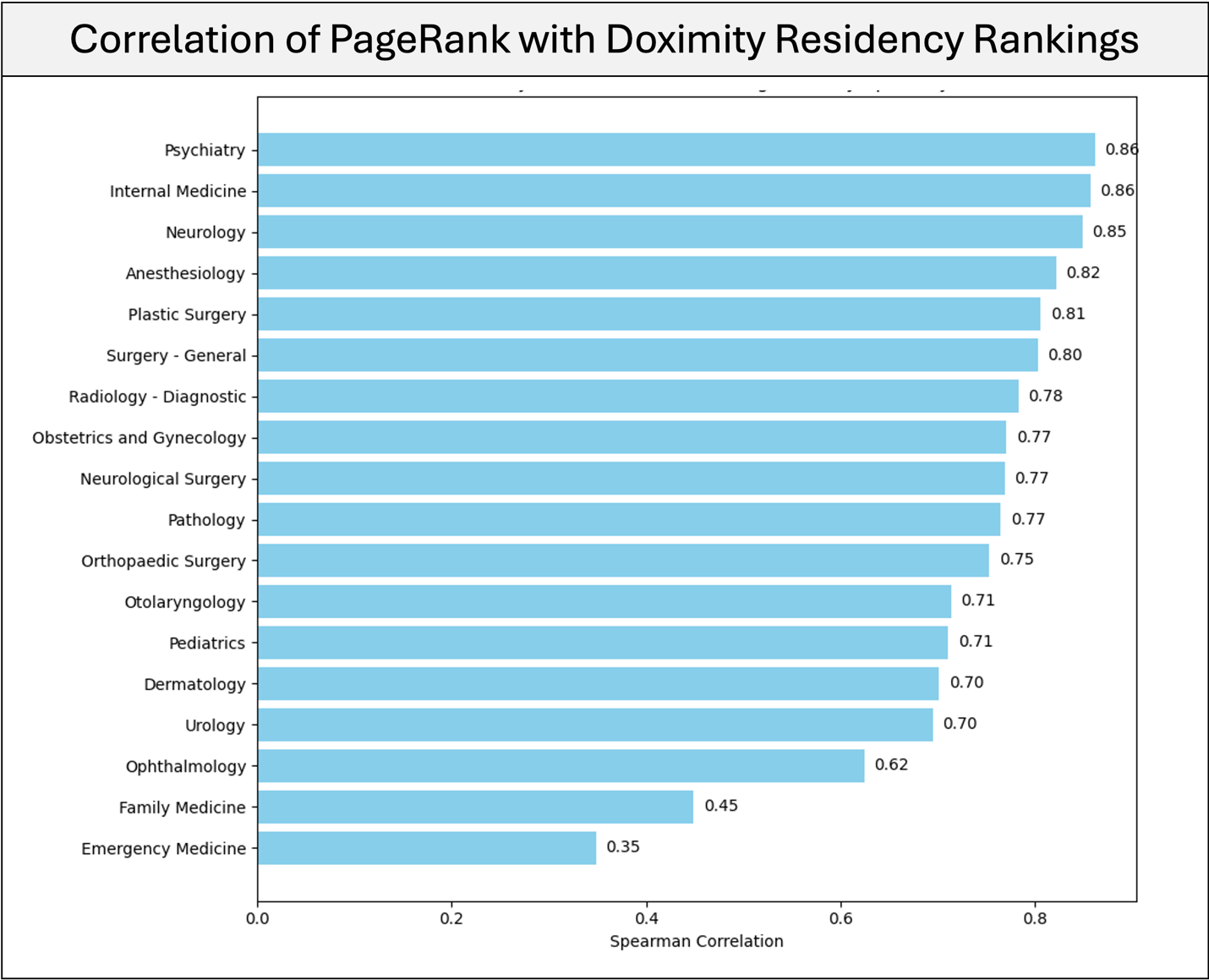

**eFigure 3. Correlation of PageRank with Doximity residency specialty rankings.** Spearman correlations between PageRank-derived institutional rankings and Doximity residency reputation rankings across specialties. PageRank showed strong concordance with survey-based rankings in most specialties.

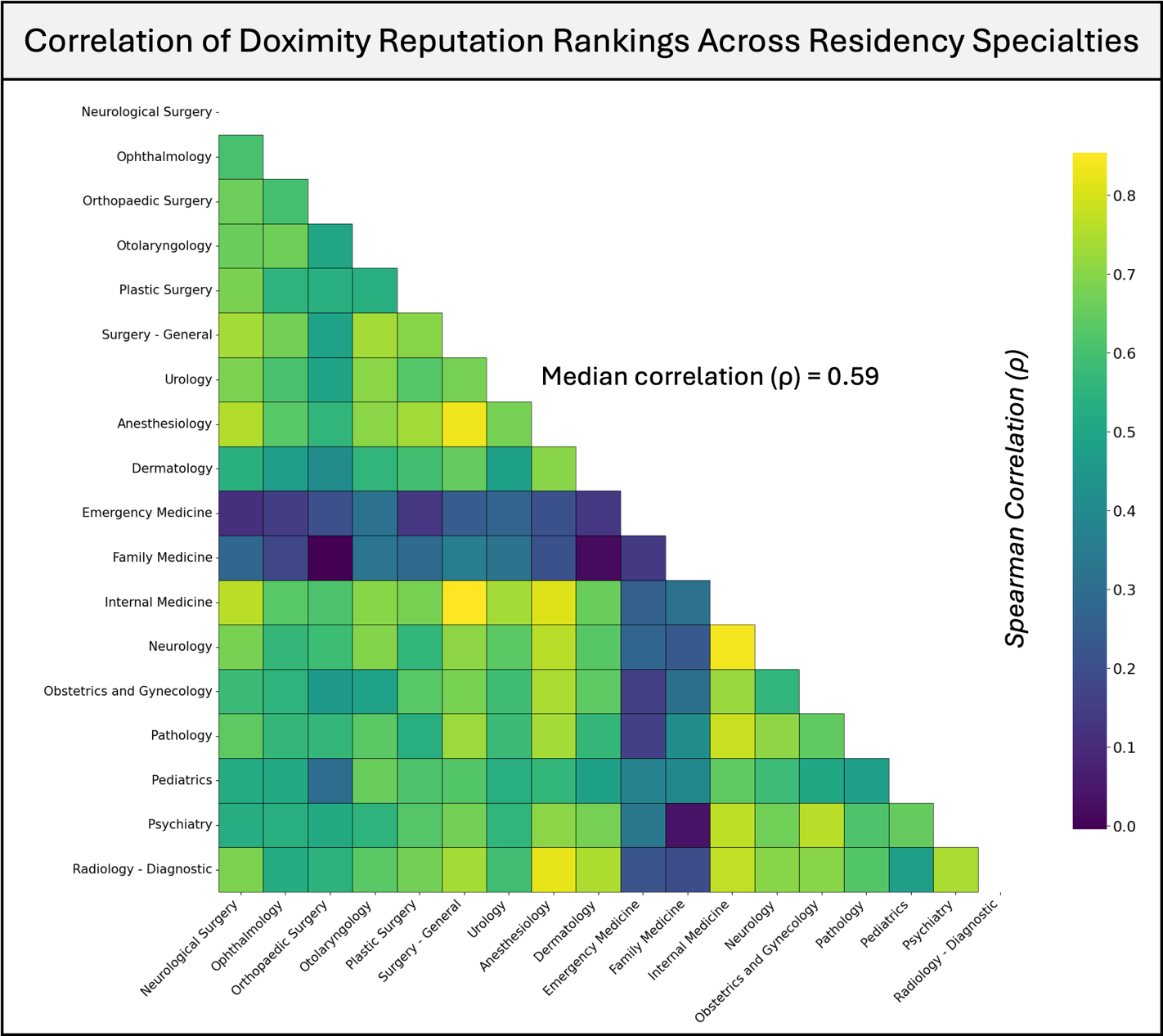

**eFigure 4**: **Spearman correlation matrix of Doximity Rankings across specialties.** Ordinal rankings of residency programs via doximity reputation surveys are similar across specialties, with a median Spearman correlation of ρ = 0.59. Notably, family medicine and emergency medicine reputation rankings diverge from the overall trend, exhibiting markedly lower correlations with other specialties.

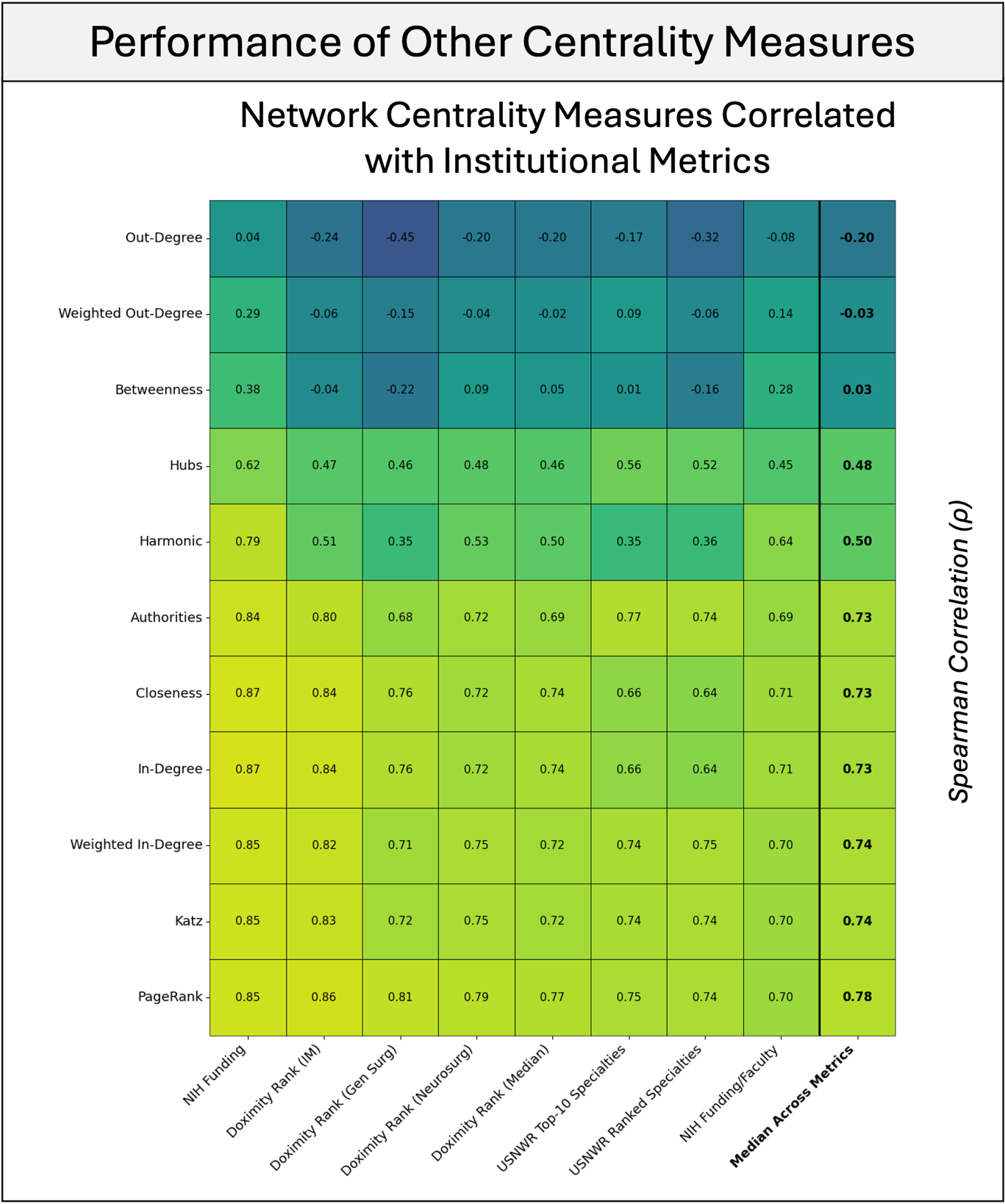

**eFigure 5. Performance of alternative network centrality measures.**
Correlations between centrality measures (PageRank, Katz, in-degree, closeness, HITS authorities and hubs, among others) and external indicators of institutional standing, including NIH research funding, Doximity residency reputation rankings, and U.S. News specialty rankings. While several measures show moderate to strong associations, PageRank demonstrates the most consistent performance across metrics, with the highest median correlation (Spearman ρ = 0.78).

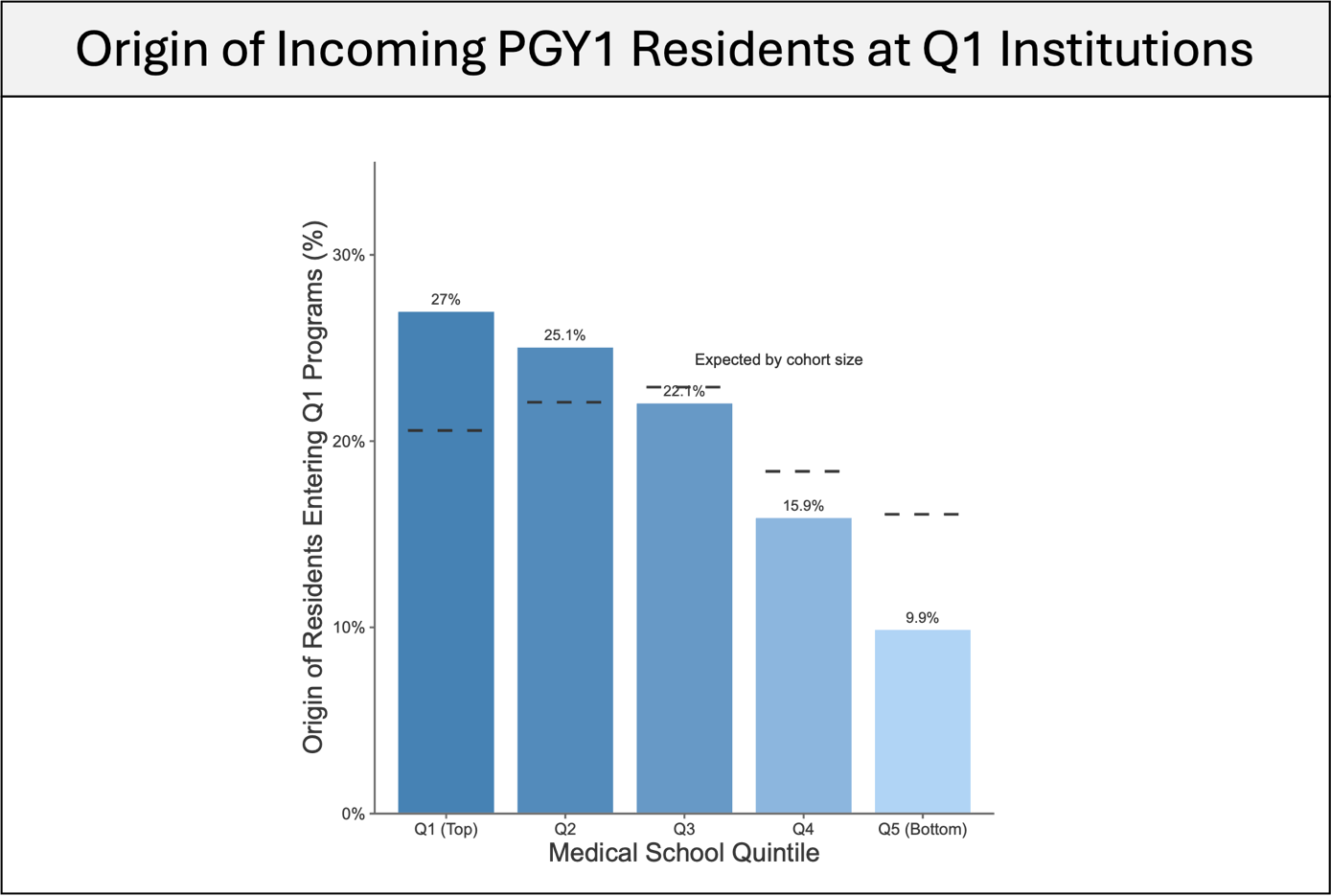

**eFigure 6. Origin of incoming PGY1 residents at top-quintile residency programs.**
Distribution of medical school PageRank quintiles among trainees entering Q1 (top-quintile) residency programs. Q1 programs recruit residents from all quintiles; however, graduates from higher-ranked institutions (Q1–Q2) are overrepresented relative to cohort size, whereas those from lower-ranked institutions (Q3–Q5) are underrepresented, most notably for Q5.

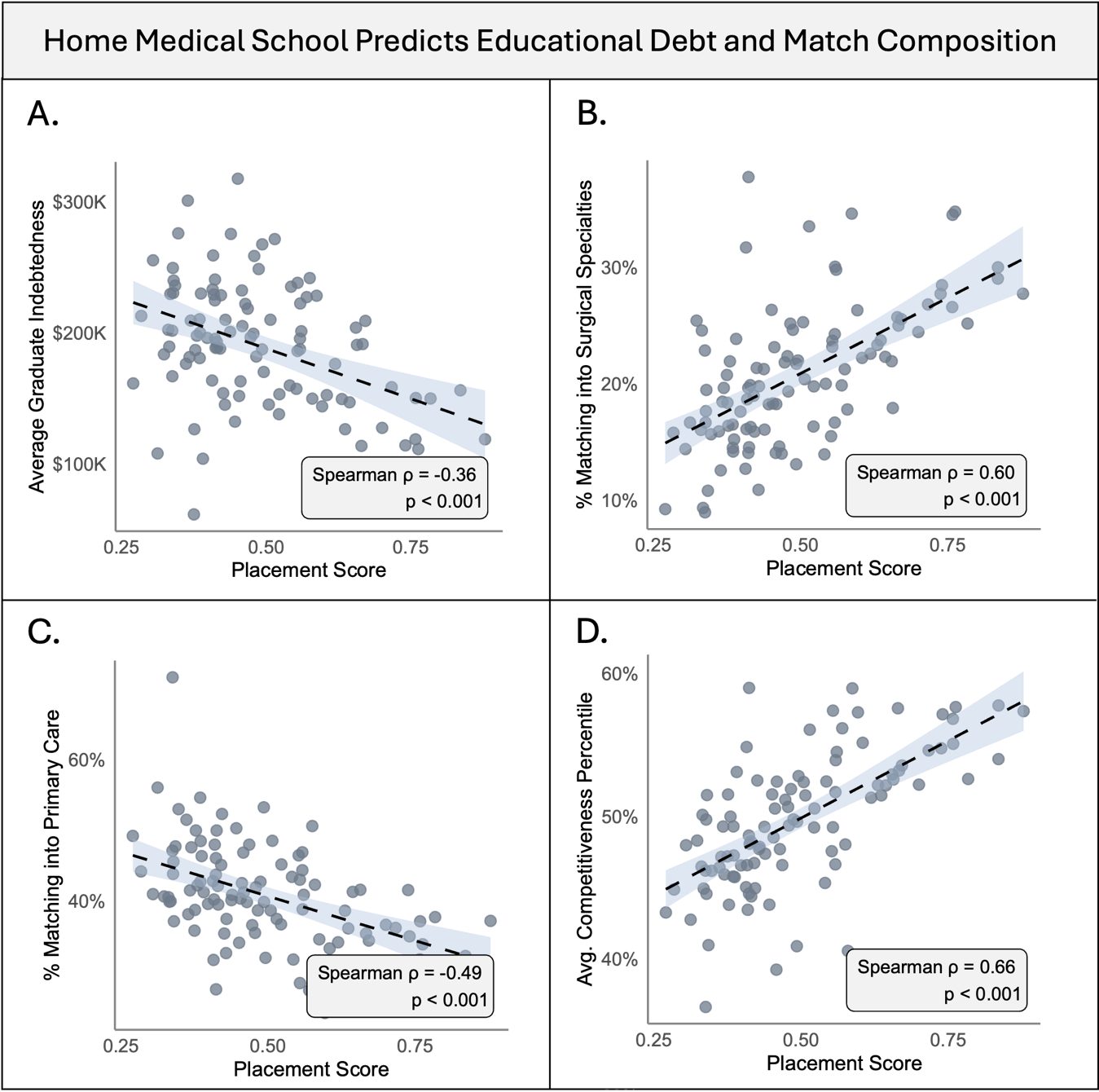

**eFigure 7. Institutional placement score predicts debt and specialty composition.**
Each point represents a U.S. MD-granting institution. Placement score, defined as the median PageRank percentile of residency destinations for a school’s graduating class, was associated with multiple student outcomes. Using Spearman rank correlation, higher placement score was associated with lower average graduate indebtedness (**A**), greater entry into surgical specialties (**B**), lower entry into primary care (**C**), and higher average specialty competitiveness (**D**; defined using specialty-specific unmatched rates among U.S. MD seniors).

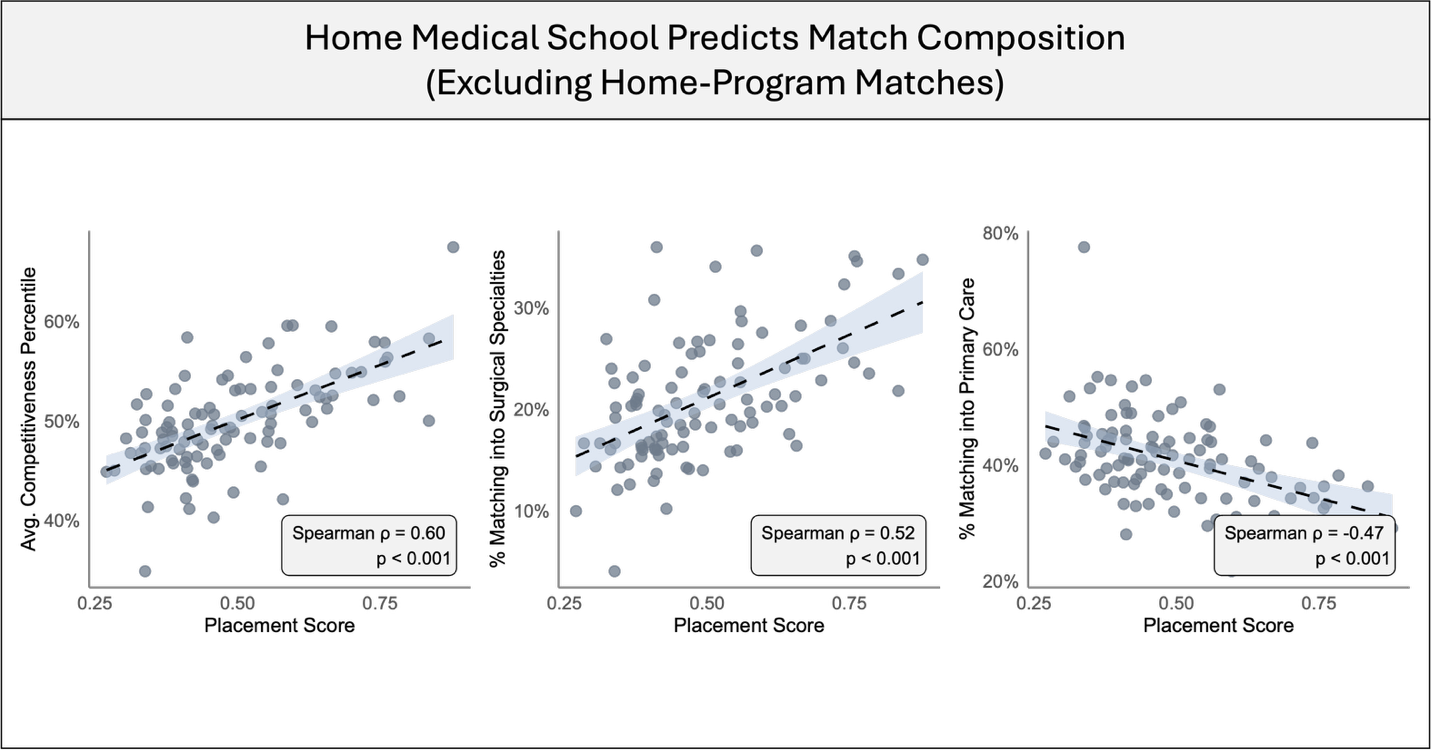

**eFigure 8. Placement score and specialty composition after excluding home-program matches.**
Each point represents a MD-granting institution. Associations between medical school placement score and specialty composition after removing matches to a student’s home institution. Higher placement score remained associated with greater entry into competitive and surgical specialties and lower entry into primary care, indicating that these relationships are not explained by preferential matching into home programs.

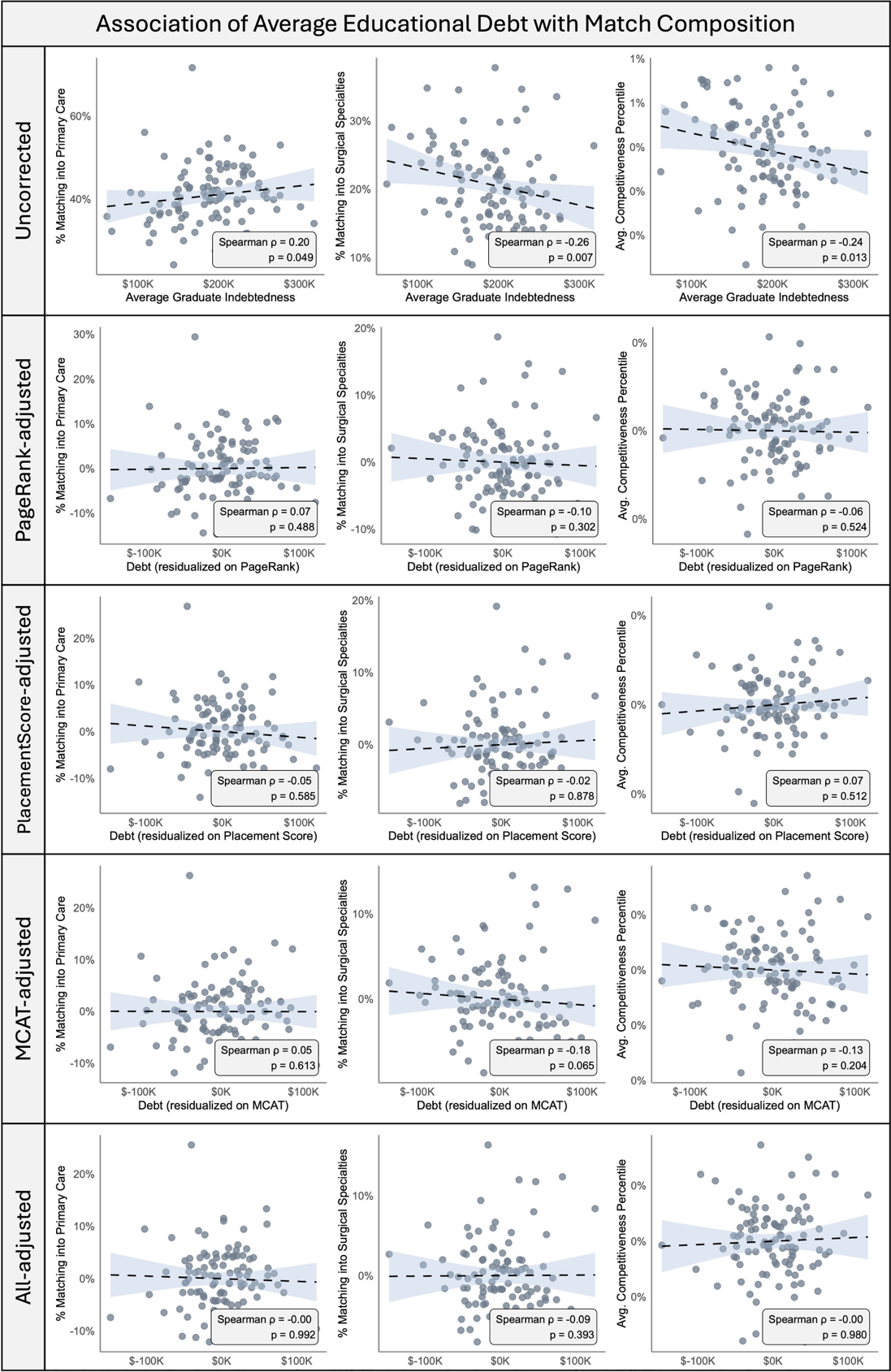

**eFigure 9. Association of educational debt with specialty composition before and after accounting for network position.**
Each point represents a MD-granting institution. In unadjusted analyses (top row), higher mean graduate indebtedness was associated with lower average specialty competitiveness, reduced entry into surgical specialties, and greater entry into primary care. After adjusting for institutional characteristics (bottom row), associations were no longer observed.

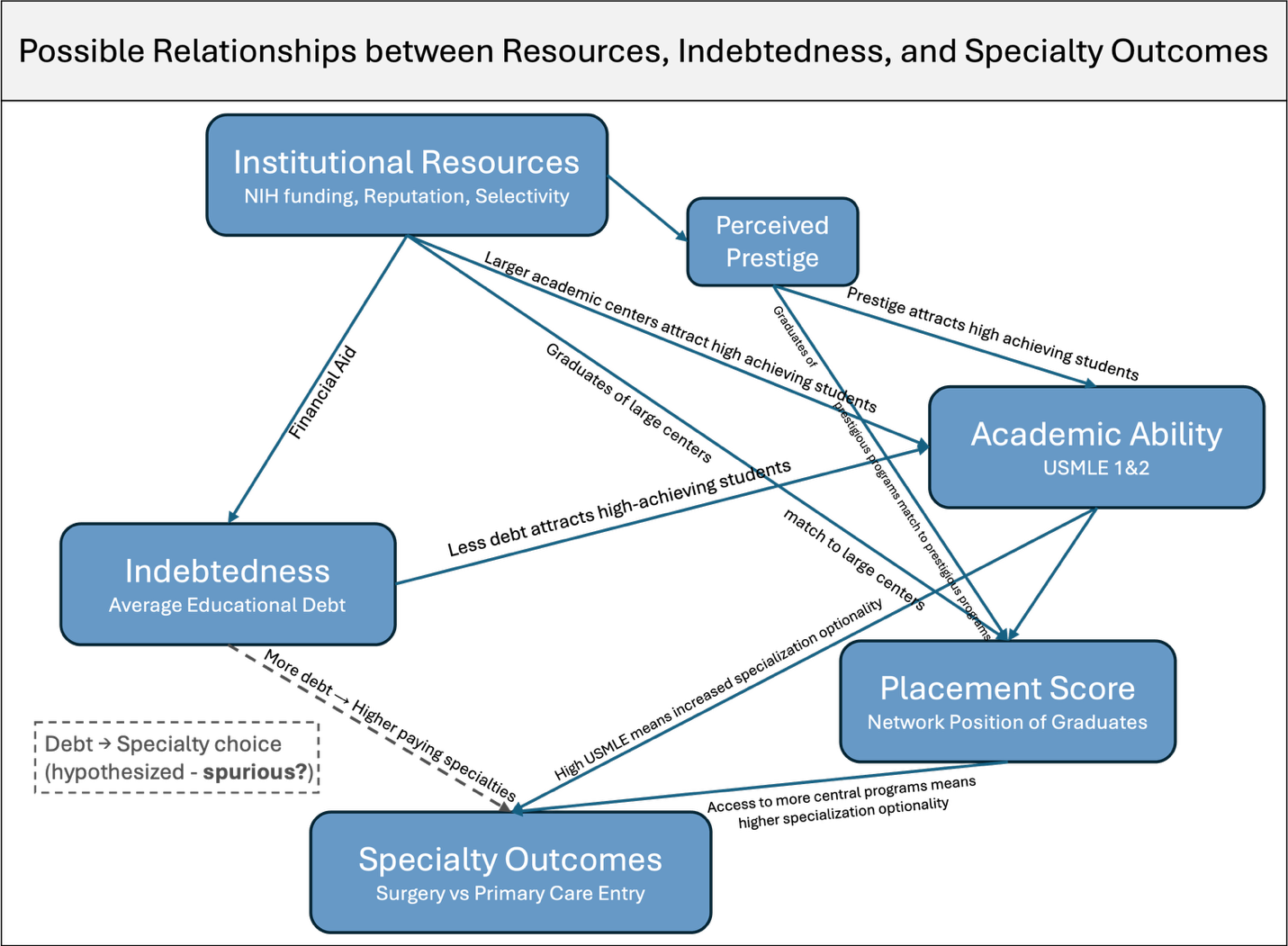

**eFigure 10. Proposed causal framework linking institutional resources, graduate indebtedness, and specialty outcomes.**
Directed acyclic graph (DAG) illustrating hypothesized relationships between institutional resources, graduate indebtedness (average educational debt), academic ability, placement score (network-derived position of matched residency programs), and specialty outcomes. Institutional resources are modeled as a common upstream factor influencing both indebtedness and access to more central residency programs. Placement score and academic ability are hypothesized to drive specialty outcomes, while the direct effect of indebtedness is uncertain (dashed line), potentially reflecting confounding by shared upstream factors.
